# Advantages of a Two-Stage Randomized Trial Design to Evaluate Antimicrobial Treatment Strategies: a Simulation Study

**DOI:** 10.64898/2026.03.16.26347803

**Authors:** Juan Gago, Christopher Boyer, Marc Lipsitch

**Affiliations:** Department of Epidemiology, Harvard T.H. Chan School of Public Health, Boston, MA, USA; Department of Medicine, Case Western Reserve School of Medicine, Cleveland, OH, USA; Department of Quantitative Health Sciences, Cleveland Clinic, Cleveland, OH, USA; Department of Medicine (Division of Infectious Diseases), Stanford University School of Medicine; Department of Biology, Stanford University; Freeman Spogli Institute for International Studies (Center for International Security and Cooperation, CISAC), Stanford, CA, USA

**Keywords:** Causal inference, interference, drug resistance, antimicrobials, agent-based models, study design

## Abstract

**Background:** Antimicrobial prescribing policies affect not only treated patients but also their contacts. Two-stage randomized (2SR) designs can be used to estimate these spillover effects, yet this study design has not been widely applied to evaluate antimicrobial strategies.

**Methods:** We developed a stochastic agent-based model that simulates a hospital ward with two competing bacterial strains (drug-A-susceptible and drug-A-resistant). We used the simulation to emulate a 2SR trial: six hospital ward clusters were randomized 1:1 to either a 90/10 (90% Drug A, 10% Drug B, drug B was assumed to have no known resistance) or 50/50 treatment allocation strategy; individuals within clusters were then randomized to Drug A or Drug B following the assigned cluster-level allocation strategy. We estimated direct, indirect, total, and overall causal effects on incident infection and mortality. Sensitivity analyses varied the treatment effect, transmission rate, mortality structure, and number of clusters.

**Results:** The direct effect of drug choice showed that Drug A recipients had higher mortality (due to non-concordant treatment of resistant infections). This effect varied over time as the ward’s strain ecology diverged between strategies. There was also an indirect effect for Drug A recipients—reflecting spillover from higher resistant-strain prevalence under 90/10—but it was approximately null for Drug B recipients, whose broad-spectrum coverage insulated them from changes in the ward strain distribution. The overall effect—the policy-level comparison—showed that the 50/50 strategy reduced total mortality, but this net benefit concealed a redistribution: resistant-strain deaths decreased while susceptible-strain deaths increased, a consequence captured by the overall effect but invisible to the direct effect. These findings were qualitatively consistent across all sensitivity scenarios.

**Conclusions:** We demonstrate that antimicrobial prescribing produces spillover effects not captured by conventional individually randomized trials. These effects can substantially alter treatment outcomes in a population. We propose that the 2SR design, grounded in a formal causal framework for interference, is better suited for evaluating population-level effects of antimicrobial strategies—whether implemented as a randomized trial or emulated with observational data.

## Introduction

Antimicrobial resistance (AMR) threatens to undermine gains of modern medicine by reducing treatment effectiveness.^1^ The challenge for clinicians and policymakers is to identify antimicrobial strategies that achieve two goals simultaneously: treating infections effectively in the individual patient and limiting the emergence and spread of resistance in the population.

Most of the evidence on antimicrobial therapies comes from individually randomized trials. These trials estimate the causal effect of treatment on treated patients but provide little information about spillover effects—how the treatment of one patient may affect the outcomes of others (e.g. patients’ contacts in the same ward).^2^ Ignoring spillover effects can misrepresent the consequences of antimicrobial treatment strategies.^3^

Two-stage randomized (2SR) designs provide a framework for simultaneously estimating direct and indirect effects.^2^ Clusters (e.g., hospital wards) are first randomized to different levels of treatment coverage, and then individuals within clusters are randomized to treatment strategies consistent with the cluster assignment. Such designs have been applied to vaccines and HIV prevention interventions, where indirect effects are expected to be substantial,^4^ and recent work has formalized their connection to the target trial framework. ^5^ To our knowledge, no applications have been developed for antimicrobial treatment strategies.

A few cluster-randomized antimicrobial trials exist. Van Duijn et al. ^6^ conducted a cluster-randomized crossover trial of antibiotic cycling versus mixing across European ICUs, but the design compared ward-level policies without individual-level randomization, precluding separation of direct and indirect effects. Arzika et al.^7^ analyzed spillover in the AVENIR trial of mass azithromycin distribution in Niger, finding that treating older siblings reduced infant mortality beyond direct protection—the first antimicrobial trial to formally estimate spillover on survival. However, neither trial adopted the two-stage structure needed to simultaneously identify both direct and indirect components. Agent-based models (ABMs) have explored related questions, such as the evaluation of carbapenem-resistant *Enterobacterales* decolonization strategies,^8^ but they have not been structured to emulate 2SR designs for antimicrobials.

In this paper, we extend the 2SR framework to antimicrobial strategies by implementing it in an ABM. We define clusters as hospital wards and randomize both cluster-level coverage (the proportion of patients treated with Drug A, which covers only susceptible infections, versus Drug B, which covers both susceptible and resistant infections) and individual-level treatment assignment (which specific drug a patient receives). The ABM serves as the data-generating mechanism from which the target 2SR trial is emulated, allowing us to quantify direct, indirect, and overall effects of antimicrobial prescribing policies under controlled conditions. The model is intentionally simplified to evaluate and assess the spillover mechanism; different parameter settings could be tested to correspond to particular clinical settings.

## Methods

### Model Setup

We developed a discrete-time, stochastic ABM to simulate a 2SR trial of antimicrobial treatment strategies in a hospital setting. Each agent represents an individual patient followed over 350 timesteps within a simulated ward structure. We used R software version 4.4.1 and the ABM package (version 0.4.3)^9^ for simulations.

Each agent occupies one of the following mutually exclusive health states: uninfected (*X*); colonized with a drug-susceptible (*S*) or drug-resistant (*R*) strain; symptomatic and treated with Drug A or Drug B (*TS*_*A*_, *TS*_*B*_, *TR*_*A*_, *TR*_*B*_); symptomatic and untreated (*US, UR*); discharged alive (*Z*); or deceased (*D*). The state-transition diagram is provided in eFigure 1.

#### Admission and Discharge

Admissions follow a Poisson process at rate *λ* = 10. New admissions enter state *X* (uninfected) with probability *m* = 0.75, or state *S* (susceptible infection) with probability 1 − *m*. No new admissions arrive with resistant infections; all resistance in the model arises from within-ward transmission. Discharge (*Z*) and death (*D*) transitions apply only to symptomatic or treated agents, at rates *µ* = 0.095 and *δ* = 0.025 (with state-specific multipliers), respectively.

#### Transmission

Homogeneous mixing within wards leads to:

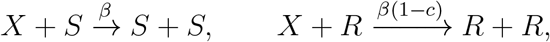

where *β* = 0.4 is the transmission rate and *c* = 0.01 is the fitness cost of resistance.

#### Symptom Onset and Treatment

Symptom onset occurs at rate *σ* = 0.5 (mean 2 timesteps to clinical manifestation). Symptomatic agents enter treatment with probability *ρ* = 0.9, receiving Drug A with cluster-specific probability *π* and Drug B with probability 1 − *π*. Untreated symptomatic patients remain in *US* or *UR*.

#### Recovery

Recovery transitions are:

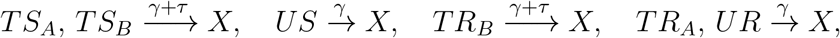

with *γ* = 0.20 and treatment effect *τ* = 0.7. The treatment effect accelerates recovery for concordant therapy: both drugs cover susceptible infections, but only Drug B covers resistant infections. Drug A recipients with resistant infections (*TR*_*A*_) recover at the untreated rate. Note that Drug B is modeled as fully effective against both strain types; in practice, broad-spectrum agents may have reduced efficacy against some resistant strains, which would attenuate Drug B’s advantage. We adopt this simplification to isolate the spillover mechanism.

#### Mortality Rates by Stratum

The primary outcome is death. The baseline death rate *δ* = 0.025 is modified by state-specific multipliers reflecting clinical severity: concordant susceptible treatment (*TS*_*A*_, *TS*_*B*_): 1 × *δ*; concordant resistant treatment (*TR*_*B*_): 1.5×*δ*; untreated susceptible (*US*): 2×*δ*; discordant resistant treatment (*TR*_*A*_): 2.5 × *δ*; and untreated resistant (*UR*): 3 × *δ*. Thus mortality increases with both lack of treatment and discordance between the infecting strain and the drug administered: concordant therapy is best, followed by concordant therapy for a harder-to-treat resistant strain, then absence of any treatment, then treatment with the wrong drug against a resistant strain, and finally no treatment of a resistant infection. Sensitivity analyses include a uniform-mortality scenario (1 × *δ* for all states) to assess the impact of this mortality structure.

#### Simulation Procedure

We initialized 1,000 agents (50 in *S*, 50 in *R*, remainder in *X*) and simulated 6 clusters: 3 assigned to *π* = 0.9 (90/10 strategy, *α*_1_) and 3 to *π* = 0.5 (50/50 strategy, *α*_0_). Each cluster was replicated 100 times to capture stochastic variability, yielding 600 total simulation runs.

### Causal Framework

We define causal effects within the potential outcomes framework under the assumption of *partial interference* ^2,10^ and *stratified interference*.^2^ Let 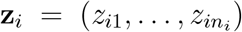 denote the treatment vector for cluster *i*, where *z*_*ij*_ ∈ {*A, B*}. The potential outcome for individual *j* in cluster *i* is *Y*_*ij*_(**z**_*i*_).

Under partial interference, an individual’s outcome may depend on treatments assigned to others in their cluster, but not on treatments in other clusters:

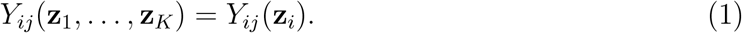

Under stratified interference, *Y*_*ij*_(**z**_*i*_) depends on **z**_*i*_ only through (a) the individual’s own treatment *z*_*ij*_ and (b) the allocation strategy *α*(**z**_*i*_). These assumptions relax the standard Stable Unit Treatment Value Assumption (SUTVA) and enable us to define individual average potential outcomes 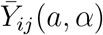, where the bar denotes an average over all treatment vectors consistent with strategy *α* and own treatment *a*.

We denote the population average as 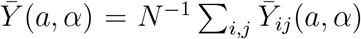, suppressing individual subscripts, and 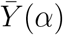 as the marginal average over treatment arms under strategy *α*.

Following Hudgens and Halloran, ^2^ we define:

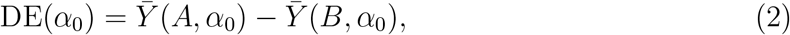

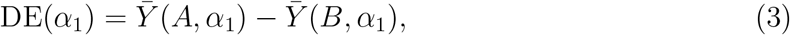

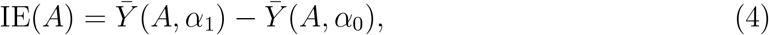

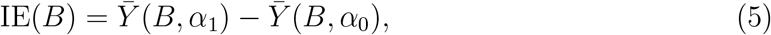

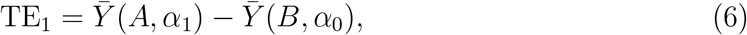

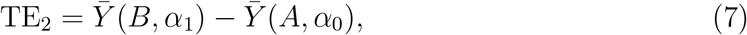

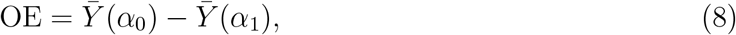

where *α*_0_ denotes the 50/50 strategy and *α*_1_ denotes the 90/10 strategy. Following Hudgens and Halloran, ^2^ the DE is the *direct effect* : the causal effect of an individual’s own treatment assignment, holding the cluster-level allocation strategy fixed. The IE is the *indirect effect* : the causal effect of changing the allocation strategy (i.e., the treatments assigned to others in the cluster) while holding the individual’s own treatment fixed. The IE is nonzero only if interference between individuals is present—that is, only if one patient’s treatment assignment affects another patient’s outcome. The TE_1_ is the *total effect*, a composite contrast that simultaneously changes both individual treatment and cluster strategy; it decomposes as TE_1_ = DE(*α*_1_) + IE(*B*) = IE(*A*) + DE(*α*_0_). The OE is the *overall effect* : the policy-level comparison of outcomes under two allocation strategies, marginalized over individual treatment assignments. Positive values indicate higher mortality; negative OE indicates that the 50/50 strategy reduces mortality.

Because both Drug A and Drug B are active treatments, a second total effect arises (eFigure 2): 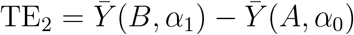, comparing Drug B recipients under 90/10 (where resistant strains predominate) with Drug A recipients under 50/50 (where resistance is suppressed). It decomposes as TE_2_ = IE(*A*) − DE(*α*_1_) = IE(*B*) − DE(*α*_0_). In vaccine trials with a placebo arm, only one total effect is defined (vaccinated under high coverage vs. unvaccinated under low coverage). With two active treatments, TE_2_ answers whether Drug B’s broad-spectrum coverage compensates for being in a high-resistance ward—a question that does not arise when one arm is inert.

We use *spillover* to refer broadly to any consequence of one patient’s treatment on another patient’s outcome—the general phenomenon that arises when interference is present. The indirect effect is one formal measure of spillover, but the overall effect also captures spillover, since it compares two allocation strategies whose outcomes differ precisely because of interference. Thus, “indirect effect” refers specifically to the IE estimand, while “spillover” describes the broader phenomenon that the 2SR design is intended to detect.

Although the Hudgens–Halloran decomposition was developed for vaccine trials comparing vaccination to placebo,^2^ the framework extends to any two-treatment 2SR design under partial interference.^10^ The estimands remain well-defined; what changes is their interpretation. In vaccine studies, the DE captures individual level benefit of vaccination at a given community vaccination level and the IE captures herd immunity. In our antimicrobial application, the DE captures the effect of drug choice (narrow-versus broad-spectrum), and the IE captures the spillover effect of the allocation strategy on same-drug recipients through altered resistance dynamics—a mechanism qualitatively distinct from herd immunity.

Because both cluster-level strategy and individual treatment are randomized, exchangeability, consistency, and positivity hold by design. When emulating this design with observational data, confounding adjustment at both levels would be essential.^11^

In our simulation, *Y*_*ij*_ = 1 if agent *j* in cluster *i* dies during the 350 timesteps, and 0 otherwise. The allocation strategy *α* corresponds to the cluster-level Drug A probability *π*, and individual treatment *z*_*ij*_ is realized through Bernoulli randomization at symptom onset. The potential outcomes *Y*_*ij*_(**z**_*i*_) are generated by the ABM’s transmission, treatment, and mortality dynamics; the estimands above are computed by averaging realized outcomes across agents and replicates.

#### Inference and Simulation Intervals

Point estimates were computed by averaging outcomes across replicates. Ninety-five percent simulation intervals (SI) were computed as empirical 2.5th–97.5th percentiles across 100 replicates per cluster-strategy combination. SIs reflect stochastic variability across simulation replicates.

##### Note on sign convention

All estimands are defined such that positive values indicate higher mortality in the first term. The OE is defined as *α*_0_ minus *α*_1_, so a negative OE indicates that the 50/50 strategy reduces mortality.

### Sensitivity Analyses

To assess whether the 2SR design can detect and quantify spillover under different model specifications, we conducted one-way sensitivity analyses varying nine dimensions independently: (1) treatment effect magnitude (*τ* ∈ {0.3, 0.7, 1.0, 1.5}); (2) transmission rate (*β* ∈ {0.2, 0.4, 0.6, 0.8}); (3) fitness cost of resistance (*c* ∈ {0.01, 0.10, 0.20}); (4) symptom onset rate (*σ* ∈ {0.3, 0.5, 1.0}); (5) mortality structure (uniform *δ* = 0.025 across all states vs. differential); (6) treatment probability (*ρ* ∈ {0.5, 0.9, 1.0}); (7) proportion admitted uninfected (*m* ∈ {0.50, 0.75, 0.90}); (8) initial conditions (*S*_0_/*R*_0_: 100/50, 50/100, 25/25 vs. baseline 50/50); and (9) cluster configuration (3 vs. 6 clusters per arm). We also varied the allocation strategies: the high-Drug-A arm (*π*) was set to 0.60–0.95 and the low-Drug-A arm to 0.30–0.70, compared with the baseline 90/10 vs. 50/50. The purpose of these analyses is to verify that the qualitative findings—positive DE, positive IE for Drug A recipients, negative OE, and the competitive exclusion tradeoff—hold under a range of plausible parameter configurations. Full results are reported in eTable 1.

## Results

The simulation comprised 6 hospital wards, with 3 randomized to the 90/10 strategy and 3 to the 50/50 strategy. Each ward was initialized with 1,000 agents, with mean admissions of 10 per timestep and discharge and death rates approximately balancing the admission flow. Under the 50/50 strategy, resistant strains were suppressed while susceptible infections dominated; under the 90/10 strategy, resistant strains persisted at higher prevalence. All point estimates reported below are averages across simulation replicates; simulation intervals indicate stochastic variability.

### Causal Effects

Table 2 presents the causal effect estimates. The average direct effect of Drug A versus Drug B was positive under both strategies (DE(*α*_0_) = +3.66 percentage points [pp]; DE(*α*_1_) = +4.99 pp), indicating higher mortality among Drug A recipients. This is expected: Drug A does not cover resistant infections, so patients with resistant infections treated with Drug A face the elevated *TR*_*A*_ mortality rate (2.5*δ* = 0.0625), while Drug B covers both strain types. The direct effect is larger under the 90/10 strategy, where greater Drug A use means a larger fraction of resistant-infected patients receive non-concordant therapy.

**Table 1:** Model parameters.

| Parameter | Symbol | Value | Justification |
| --- | --- | --- | --- |
| Admission rate | $\lambda$ | 10 | Maintains $\approx 1000$ agents |
| Prob. uninfected admission | $m$ | 0.75 | Baseline prevalence |
| Initial S/R infected | $S_0, R_0$ | 50 | Seeds infection pressure |
| Transmission rate | $\beta$ | 0.4 | Within-ward contact rate |
| Fitness cost of resistance | $c$ | 0.01 | Reduced R transmissibility |
| Recovery rate | $\gamma$ | 0.20 | Mean duration 5 timesteps |
| Treatment effect | $\tau$ | 0.7 | 78% reduction in infectious period |
| Symptom onset rate | $\sigma$ | 0.5 | Moderate latency (mean 2 timesteps) |
| Treatment probability | $\rho$ | 0.9 | High uptake |
| Discharge rate | $\mu$ | 0.095 | Balances admissions |
| Baseline death rate | $\delta$ | 0.025 | 2.5% baseline mortality |
| Prob. Drug A ($\alpha_1$) | $\pi$ | 0.90 | 90/10 strategy |
| Prob. Drug A ($\alpha_0$) | $\pi$ | 0.50 | 50/50 strategy |

*Note:* Sensitivity analyses varied $\tau, \beta, c, \sigma, \delta, \rho, m, S_0/R_0$ , number of clusters, and allocation strategies. See eTable 1.

**Table 2:** Causal effect estimates (percentage points) with 95% simulation intervals. *α*_0_ = 50/50 strategy; *α*_1_ = 90/10 strategy.

| Estimand | Definition | Estimate | 95% SI | Interpretation |
| --- | --- | --- | --- | --- |
| *A. Direct Effects (Drug A vs B within each strategy)* |  |  |  |  |
| DE($\alpha_0$) | $\bar{Y}(A, \alpha_0) - \bar{Y}(B, \alpha_0)$ | +3.66 | [+2.67, +4.78] | Own-treatment effect at 50/50 |
| DE($\alpha_1$) | $\bar{Y}(A, \alpha_1) - \bar{Y}(B, \alpha_1)$ | +4.99 | [+3.53, +6.45] | Own-treatment effect at 90/10 |
| *B. Indirect Effects (strategy change, holding own treatment fixed)* |  |  |  |  |
| IE(A) | $\bar{Y}(A, \alpha_1) - \bar{Y}(A, \alpha_0)$ | +1.24 | [−0.73, +3.34] | Indirect effect on A-recipients |
| IE(B) | $\bar{Y}(B, \alpha_1) - \bar{Y}(B, \alpha_0)$ | −0.09 | [−0.98, +0.78] | Indirect effect on B-recipients |
| *C. Total and Overall Effects* |  |  |  |  |
| TE<sub>1</sub> | $\bar{Y}(A, \alpha_1) - \bar{Y}(B, \alpha_0)$ | +4.89 | [+3.39, +6.75] | DE($\alpha_1$) + IE(B) |
| TE<sub>2</sub> | $\bar{Y}(B, \alpha_1) - \bar{Y}(A, \alpha_0)$ | −3.75 | [−5.16, −2.44] | IE(A) − DE($\alpha_1$) |
| OE | $\bar{Y}(\alpha_0) - \bar{Y}(\alpha_1)$ | −2.55 | [−4.42, −0.96] | Policy-level effect |

Note: Positive values = higher mortality in the first term. OE negative = 50/50 reduces mortality. SIs are empirical 2.5th–97.5th percentiles across 100 replicates.

For Drug B recipients, the indirect effect was approximately null (IE(B) = −0.09 pp; 95% SI: −0.98, +0.78): changing the allocation strategy had minimal impact on mortality among patients whose own treatment (Drug B) was held fixed, consistent with Drug B covering both strain types regardless of ward ecology. For Drug A recipients, the indirect effect was positive (IE(A) = +1.24 pp; 95% SI: −0.73, +3.34): under the 90/10 strategy, the ward environment has higher resistant-strain prevalence, and since Drug A does not cover resistant infections, the shift in case mix toward non-concordant treatment raises mortality among A-recipients even though their own treatment assignment is unchanged. Although the simulation interval includes the null in this baseline configuration, this reflects the limited number of clusters (6 total); the sensitivity analyses show that IE(A) increases substantially under higher transmission, higher treatment probability, and wider allocation contrasts—conditions that are plausible in real hospital settings (see Sensitivity Analyses). These patterns are visible in Figure 1, where panel A shows the 90/10 strategy producing substantially more resistant-strain deaths, while panel B reveals that even susceptible-strain deaths diverge between strategies, reflecting indirect effects operating through altered transmission dynamics.

**Figure 1:**
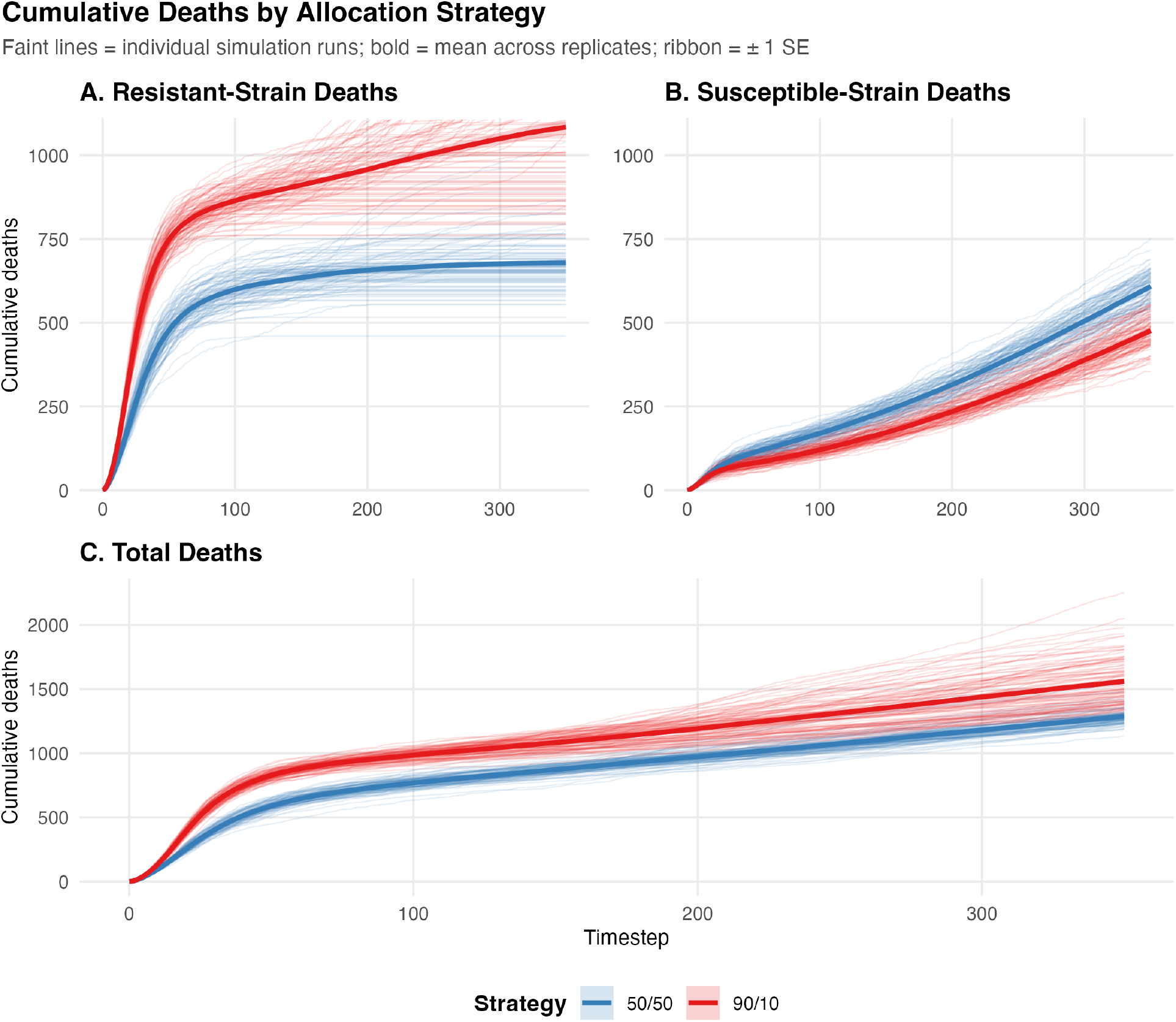
Cumulative deaths by allocation strategy. (A) Deaths from resistant infections: the 90/10 strategy produces substantially more resistant-strain deaths. (B) Deaths from susceptible infections: the 50/50 strategy produces modestly more susceptible-strain deaths, reflecting competitive exclusion. (C) Total deaths: the 50/50 strategy yields lower overall mortality. Faint lines show individual simulation runs; bold lines show means across 100 replicates; ribbons indicate ±1 SE.

The first total effect was positive (TE_1_ = +4.89 pp): Drug A recipients under 90/10 had higher mortality than Drug B recipients under 50/50. The second total effect was negative (TE_2_ = −3.75 pp; 95% SI: −5.16, −2.44): Drug B recipients under 90/10—where resistant-strain prevalence is high—still had lower mortality than Drug A recipients under 50/50, where resistance become less prevalent. This reflects the fact that patients receiving Drug B are largely insulated from the ward’s strain resistance distribution, since Drug B covers both susceptible and resistant infections regardless of what others receive. This insulation complicates the interpretation of conventional individually randomized trials, which compare treatments at a single fixed allocation and cannot reveal how drug efficacy depends on the broader prescribing environment. In vaccine trials, where one arm is a placebo, only one such total effect is defined; with two active treatments, both contrasts are informative. The overall effect was −2.55 pp (95% SI: −4.42, −0.96): the 50/50 strategy yielded 1,275 total deaths versus 1,545 under 90/10 (Table 3), a reduction of 270 deaths. However, this benefit was distributed unevenly by infection type. Resistant-infected patients benefited substantially from greater Drug B availability (fewer deaths: 674 vs. 1,068). Conversely, susceptible-infected patients experienced higher mortality under 50/50 (601 vs. 477 deaths), driven by higher cumulative incidence of susceptible infections when resistant strains are suppressed (Figure 2, Table 4).

**Table 3:** Cumulative infection incidence and deaths by allocation strategy.

| Strategy | Cumul. S Infec. | Cumul. R Infec. | Deaths (S) | Deaths (R) |
| --- | --- | --- | --- | --- |
| 50/50 | 15,568 | 5,599 | 601 | 674 |
| 90/10 | 12,251 | 6,257 | 477 | 1,068 |

**Figure 2:**
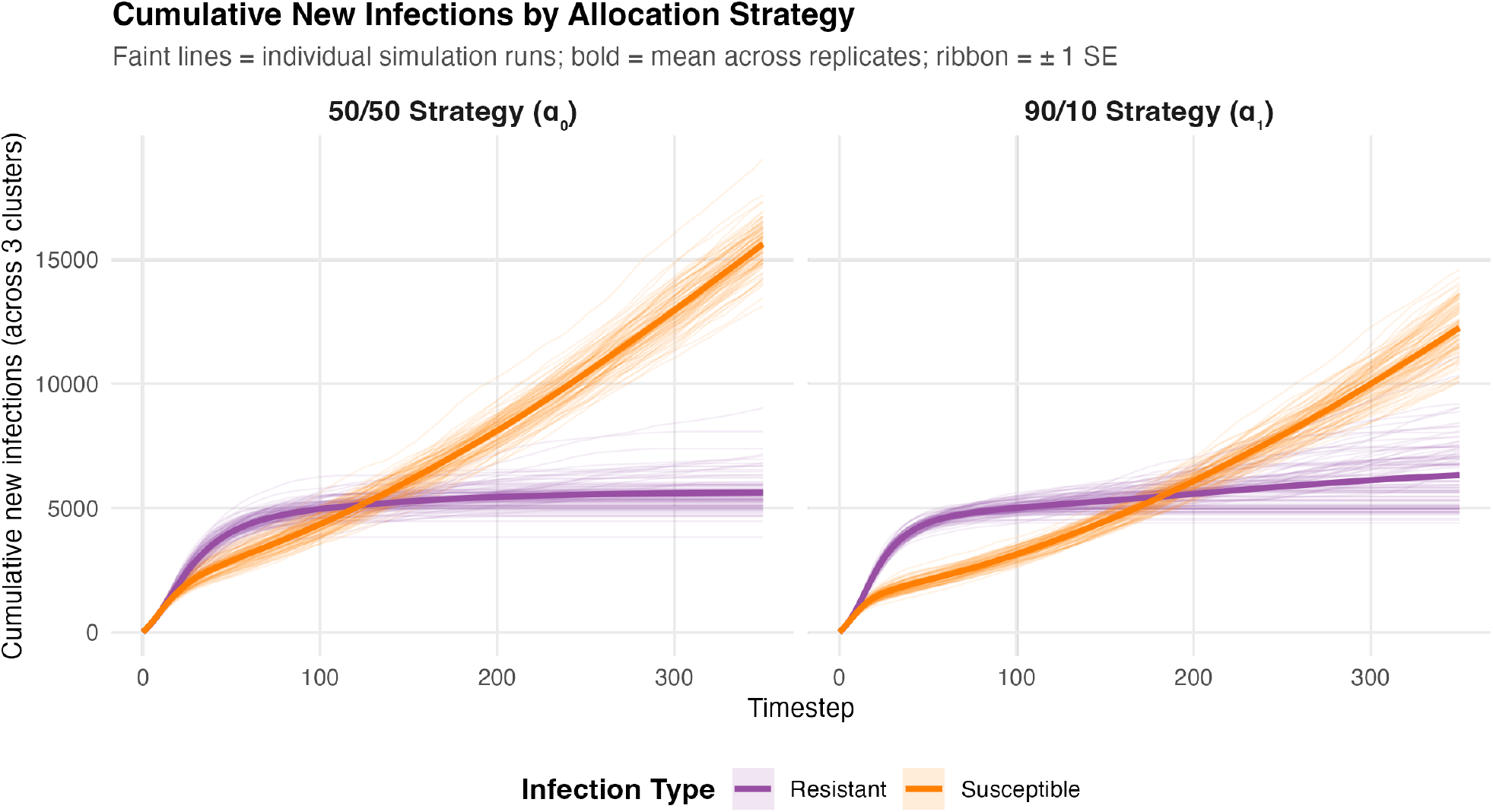
Cumulative new infections by allocation strategy and infection type. Under the 50/50 strategy, susceptible infections accumulate substantially more than under 90/10, while resistant infections are suppressed. This reversal illustrates the competitive exclusion mechanism: suppressing resistant strains with Drug B releases susceptible strains from ecological competition.

**Table 4:** Risk of death by infection type and allocation strategy, with risk differences (RD).

| Strategy | Susceptible |  | Resistant |  | RD | 95% SI |
| --- | --- | --- | --- | --- | --- | --- |
|  | Risk | SE | Risk | SE |  |  |
| 50/50 | 0.0386 | 0.0002 | 0.1200 | 0.0005 |  |  |
| 90/10 | 0.0389 | 0.0002 | 0.1710 | 0.0005 |  |  |
| RD Susceptible | | | | | +0.0004 | $[-0.005, +0.005]$ |
| RD Resistant | | | | | +0.051 | $[+0.039, +0.064]$ |

### Infection and Mortality Outcomes

Table 3 summarizes key outcomes by allocation strategy and infection type. The 50/50 strategy led to lower cumulative resistant infections (5,599 vs. 6,257) and fewer resistant-related deaths (674 vs. 1,068). Conversely, susceptible infection incidence was higher (15,568 vs. 12,251) and so was susceptible-related mortality (601 vs. 477). This redistribution is captured by the overall effect (OE = −2.55 pp): the OE reflects net mortality across both strain types, and its negative value indicates that the resistant-death reduction outweighs the susceptible-death increase. Critically, none of this redistribution is visible in the direct effect—which compares Drug A to Drug B recipients within a strategy and shows no difference for susceptible infections (DE_*S*_ ≈ 0). The mortality shift arises entirely from Drug B– intensive therapy reducing resistant strain circulation and releasing susceptible strains from ecological competition (Figure 2), a consequence of one patient’s treatment altering the strain ecology that other patients face. A standard individually randomized trial, which cannot estimate the OE, would miss this tradeoff entirely.

### Stratified Direct Effects

The marginal DE is a mixture of direct effects across infection types. To further decompose this, we computed stratified DEs conditional on whether patients had susceptible- or resistant-strain infections. Among patients with susceptible infections, the DE was near zero under both strategies (DE_*S*_(*α*_1_) = +0.17 pp; DE_*S*_(*α*_0_) = −0.04 pp), consistent with both drugs covering susceptible strains equally. Among patients with resistant infections, the DE was large (DE_*R*_(*α*_1_) = +14.75 pp, 95% SI: +12.98, +16.34; DE_*R*_(*α*_0_) = +13.93 pp, 95% SI: +12.24, +15.66): Drug A recipients with resistant infections had substantially higher mortality than Drug B recipients, because Drug A provides no coverage. The marginal DE is thus driven almost entirely by the resistant-infection stratum, weighted by the proportion of patients in each stratum. This decomposition confirms that the DE reflects discordant therapy for resistant infections, not a general drug-quality difference.

Importantly, DE_*S*_ ≈ 0 does not imply that the allocation strategy has no consequence for patients with susceptible infections. While drug choice is irrelevant for this group (both drugs are equally effective), the *number* of patients who acquire susceptible infections is shaped by the strategy through transmission dynamics: the 50/50 strategy suppresses resistant strains and releases susceptible strains from competition, increasing susceptible-strain incidence (15,568 vs. 12,251) and susceptible-strain deaths (601 vs. 477). This harm operates entirely through the indirect and overall effects—not through the direct effect of drug choice.

Moreover, because the allocation strategy reshapes the strain mix over time, the marginal DE is inherently time-varying. Figure 3 shows that the marginal DE declines from approximately 8–10 percentage points early in the simulation to near zero under the 50/50 strategy, following the decline in resistant-strain prevalence. The stratum-specific effects remain stable throughout (DE_*S*_ ≈ 0; DE_*R*_ ≈ 14 pp; eTable 2), confirming that the change in the marginal DE reflects shifting composition rather than changing treatment efficacy. This arises because antimicrobial treatment is effectively a combined intervention of infection type and drug choice^3^. As the strategy alters which infection types patients acquire, the effective treatment changes even though drug assignment does not—rendering the DE a time-varying quantity whose per-exposure components can be isolated only through stratification.^12^ Different trajectories in the composition of susceptible and resistant infections— shaped over time by the intervention itself—will render different marginal direct effects even when per-exposure efficacy is unchanged, a consideration often overlooked when evaluating antimicrobial interventions.

**Figure 3:**
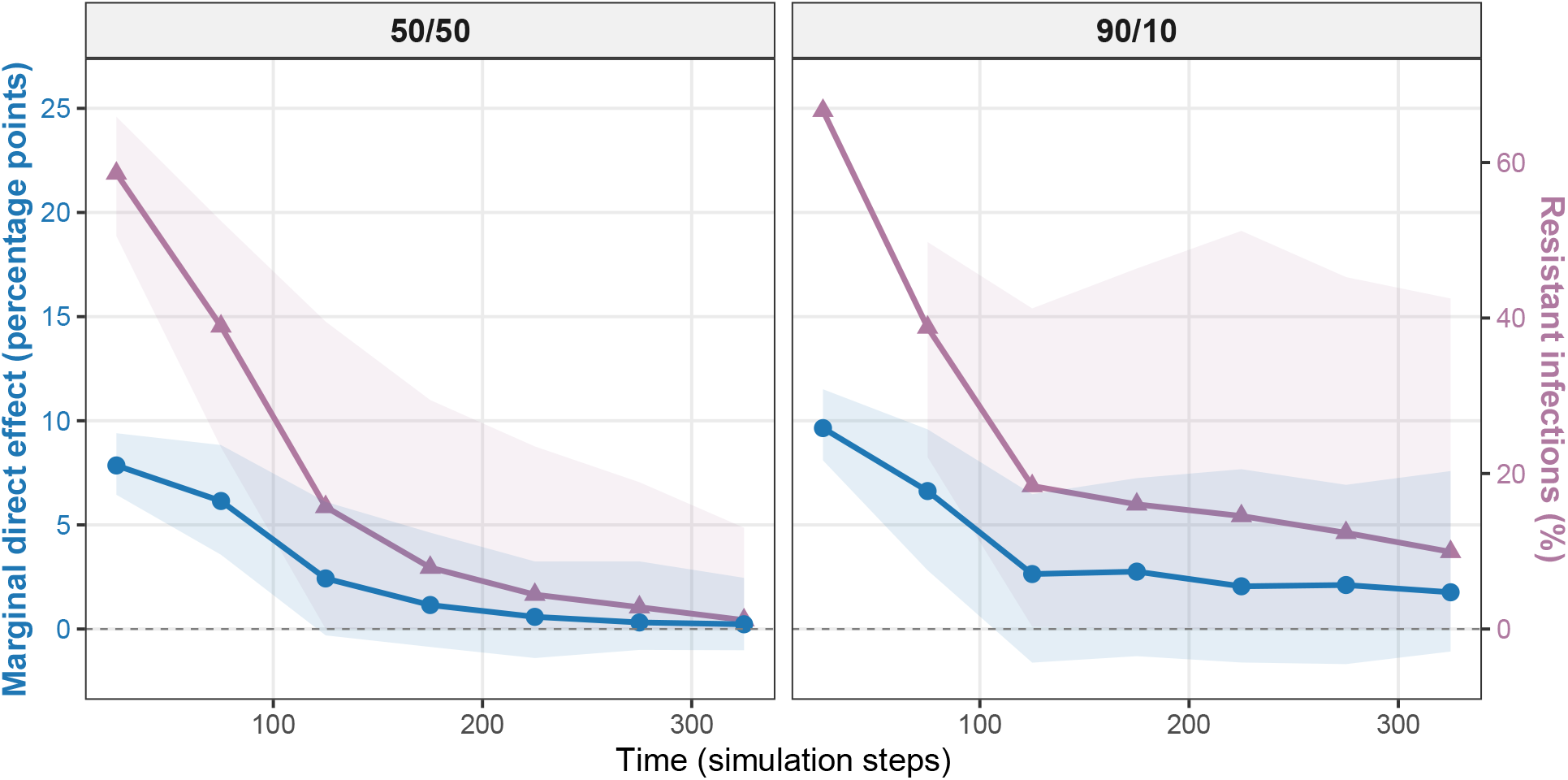
Time-varying marginal direct effect (blue circles, left axis) and proportion of treated infections that are resistant (purple triangles, right axis), by allocation strategy. The marginal DE declines over time as resistant-strain prevalence falls, reflecting the changing composition of infections rather than a change in treatment efficacy. Stratum-specific effects remain stable: DE_*S*_ ≈ 0 and DE_*R*_ ≈ 14 pp throughout (eTable 2). Under the 50/50 strategy (more Drug B), resistance is suppressed more rapidly, and the marginal DE approaches zero earlier. Shaded bands represent 95% simulation intervals across 100 replicates.

### Mechanism: Competitive Exclusion

The biological mechanism driving spillover is competitive exclusion between strains. When Drug B use is low (90/10 strategy), resistant strains are less suppressed and maintain higher prevalence, reducing the ecological niche available for susceptible-strain transmission. Conversely, when Drug B use is high (50/50 strategy), resistant prevalence declines, allowing susceptible strains to exploit the freed transmission space. This ecological dynamic is the pathway through which spillover operates: changing the allocation strategy alters strain competition, which in turn changes the infection risk that each patient faces regardless of their own treatment assignment. The indirect effect (IE) formally quantifies this pathway for patients whose own treatment is held fixed; the overall effect (OE) captures the net population-level consequence. The net effect on mortality is complex: while the 50/50 strategy prevents resistant-strain deaths, it paradoxically increases susceptible-strain incidence, which also causes mortality. Stewardship policies thus interact with transmission dynamics in non-obvious ways—a phenomenon that the two-stage design is uniquely positioned to capture.

### Sensitivity Analyses

Across all 26 sensitivity scenarios (eTables 1a–1b), the strategy with more Drug B (*α*_0_) reduced total mortality in every case (OE *<* 0; range −0.14 to −4.53 pp). However, this net benefit concealed a consistent redistribution of mortality between strain types. In every scenario, the Drug B–intensive strategy produced *more* susceptible-strain deaths than the Drug A–intensive strategy, and in all but the narrowest allocation contrast (60/40 vs. 50/50) it also reduced resistant-strain deaths (eTable 1b). In the baseline, susceptible deaths rose from 450 under *α*_1_ to 620 under *α*_0_ (+38%), while resistant deaths fell from 1,154 to 694 (−40%). This tradeoff was universal: at high transmission (*β* = 0.8), susceptible deaths increased from 448 to 839 (+87%) while resistant deaths decreased from 3,064 to 2,094 (−32%); even at low transmission (*β* = 0.2), where overall effects were attenuated (OE = −0.98 pp), susceptible deaths were modestly higher under *α*_0_ (241 vs. 233).

The magnitude of this tradeoff in mortality was most sensitive to transmission intensity, treatment efficacy, and the proportion of uninfected admissions. At *β* = 0.8, total deaths under *α*_0_ were 2,933 (839 susceptible, 2,094 resistant) versus 3,512 under *α*_1_ (448 susceptible, 3,064 resistant)—the overall benefit was large (OE = −4.53 pp), but susceptible mortality nearly doubled. When treatment efficacy was high (*τ* = 1.5), OE reached −3.79 pp; when the proportion of uninfected admissions was high (*m* = 0.90), OE was −3.27 pp, with resistant deaths 967 under *α*_0_ versus 1,291 under *α*_1_ while susceptible deaths were higher under *α*_0_ (330 vs. 258). Varying the symptom onset rate (*σ*) confirmed that results hold under both slower (*σ* = 0.3, OE = −3.82 pp) and faster (*σ* = 1.0, OE = −2.84 pp) clinical progression. Under uniform mortality or reduced treatment probability (*ρ* = 0.5), effects were attenuated but the directional tradeoff persisted.

Allocation strategy variations confirmed that the magnitude of both the overall benefit and the susceptible-death increase scaled with the contrast between strategies. The widest gap (90/10 vs. 30/70) produced a large OE (−4.00 pp) and susceptible-death gap (703 vs. 451, +56%), while the narrowest contrast (60/40 vs. 50/50) yielded OE = −0.14 pp and a modest susceptible-death difference (577 vs. 549, +5%). This gradient underscores that stewardship policies must weigh the population-level mortality reduction against the strain-specific redistribution of harm—a tradeoff that only a 2SR design can detect and quantify.

The indirect effect on Drug A recipients, IE(A), was positive in 24 of 26 scenarios and exhibited a clear dose–response pattern across several parameter dimensions. IE(A) increased with transmission intensity (+0.55 pp at *β* = 0.2; +2.67 at *β* = 0.6), treatment probability (+0.93 at *ρ* = 0.5; +1.91 at *ρ* = 1.0), treatment efficacy (+0.80 at *τ* = 0.3; +1.82 at *τ* = 1.5), and the width of the allocation contrast (−0.37 at 60/40 vs. 50/50; +2.67 at 90/10 vs. 30/70). The pattern is intuitive: conditions that amplify the ecological divergence between strategies—faster transmission, more patients treated, more effective therapy, wider prescribing differences—produce larger indirect effects. In settings with high transmission and near-universal treatment, such as intensive care units, the indirect effect on Drug A recipients would be expected to be substantially larger than in our baseline, and clearly distinguishable from zero. This underscores the practical importance of the 2SR design: precisely in the clinical settings where antimicrobial stewardship decisions matter most, the indirect effects of prescribing policy on individual outcomes are likely to be largest.

## Discussion

This simulation study demonstrates, as a proof of principle, that a 2SR design can detect and quantify spillover effects of antimicrobial prescribing strategies that would be invisible to conventional individually randomized trials. The design estimated direct effects of drug choice, indirect effects of the allocation strategy on same-drug recipients, and overall effects. Of these, the overall effect is the most policy-relevant estimand: it captures the net population-level mortality consequence of changing the prescribing strategy, including the redistribution of deaths between strain types that neither the DE nor the IE alone reveals. The DE tells us which drug is better for an individual patient; the OE tells us what happens to the ward when the prescribing policy changes. In our simulation, the OE showed that the 50/50 strategy reduced total mortality, but this benefit masked an increase in susceptible-strain deaths—a tradeoff driven entirely by ecological dynamics that would be completely undetectable in a standard individually randomized trial. The specific numerical estimates depend on model parameters and should not be interpreted as predictions for any particular clinical setting; rather, they demonstrate that the 2SR framework *can measure* these effects.

The competitive exclusion mechanism has important implications for stewardship. When broad-spectrum (Drug B) use is high, resistant strain prevalence falls. However, this releases susceptible strains from competition, leading to higher susceptible-strain incidence. While the 50/50 strategy prevents resistant-strain disease directly (fewer R-related deaths: 674 vs. 1,068), it paradoxically increases susceptible-strain deaths (601 vs. 477). This tradeoff was robust across all 26 sensitivity scenarios (eTable 1b), confirming that the redistribution of infections toward susceptible strains is an inherent consequence of broad-spectrum use. Stewardship policies changing antibiotic use practices have consequences beyond those individuals assigned to a specific treatment.^1,13^ We note that the existence of competitive relationships between resistant and susceptible strains is both consistent with ecological theory and supported by some data, but not all studies have found evidence of such competition, which may depend on the setting, bacterial species, and resistance mechanism.^14,15^

Our application extends the Hudgens–Halloran causal framework beyond its original vaccine setting, where one arm is a true control (placebo) and the indirect effect reflects herd immunity.^2,10^ In the antimicrobial context, both allocation strategies involve active treatments, so the DE compares drug choice rather than treatment versus nothing, and the IE captures how the ward-level prescribing environment influences the effect of an intervention across settings for patients whose own treatment is held fixed. This is a substantial difference: whereas vaccine-trial spillover is uniformly protective (herd immunity benefits the unvaccinated), antimicrobial spillover (operating through changes in the distribution of strains) can simultaneously harm one group while benefiting another. The 2SR framework is well suited to quantify this tradeoff.

An important feature of the antimicrobial setting is that the effects are inherently time-varying. The stratum-specific direct effects—DE_*S*_ and DE_*R*_—remain relatively stable over time (eTable 2), but the marginal DE is a time-varying mixture of these, with weights determined by the proportion of resistant infections. In our simulation, resistant-strain prevalence declines over time as competitive exclusion suppresses the resistant strain, and the marginal DE correspondingly decreases from approximately 8–10 percentage points early in the trial to near zero (Figure 3). The rate of decline is faster under the 50/50 strategy, where greater Drug B use suppresses resistance more rapidly. The IE, TE_1_, and OE similarly evolve over time as the ecological consequences of prescribing accumulate. This contrasts with vaccines used as pre-exposure prophylaxis, where the DE may be relatively stable once immunity is established. However, post-exposure prophylaxis with vaccines faces analogous complications, as the intervention’s effectiveness depends on the timing and type of exposure; ^16^ estimating outcomes averted by a vaccination program is further complicated when rollout occurs over time.^17^ The reason the antimicrobial setting is inherently time-varying is that the effective treatment a patient receives depends jointly on the drug assigned and the infection type acquired—the latter being shaped by the allocation strategy through ecological dynamics. ^3^ The drug–pathogen combination constitutes the actual intervention only in a challenge trial where both are assigned;^3^ in randomized or observational settings, the link is mediated by transmission. In our simulation, Drug A given to a patient with a susceptible infection is a qualitatively different treatment from Drug A given to a patient with a resistant infection, yet both are counted under the same arm. The stratified DE (DE_*S*_ ≈ 0; DE_*R*_ ≈ 14 pp) makes this explicit: the marginal DE is a time-varying mixture of these stratum-specific effects, with weights that shift as the ecology evolves. The per-exposure effect—the direct effect conditional on exposure type—has been proposed as a more stable estimand for infectious disease interventions, but exposure status outside of simulations or challenge trials is rarely known.^3,12^ This time-dependence also raises questions about transportability: the causal effects estimated in one setting may not generalize to settings with different baseline resistance prevalence, transmission rates, or drug availability, since these factors determine the strain mix and thus the effective intervention. Formal methods for transporting causal effects under interference remain an open area of research that we are currently pursuing.

The 2SR design should be considered for future antimicrobial trials where: (1) spillover via transmission is expected to be substantial; (2) AMR dynamics (e.g., strain competition) may alter outcomes of interest; (3) quantifying the spillover effect is a policy concern; and (4) the intervention operates at a system level (stewardship policies, prescribing guidelines). Our sensitivity analyses verify that the qualitative findings hold under a range of model specifications.

The target trial framework, combined with ABM emulation, provides a roadmap for observational emulation. One would define clusters as hospital wards, identify prescribing strategies, control for confounders at both levels (ward type, patient acuity, baseline resistance), and estimate effects using inverse probability weighting or g-computation.^11^ Electronic health records from hospital networks could support such emulation. The stratified interference assumption requires that confounding be controlled within clusters; unmeasured cross-cluster effects would violate partial interference.

This study has several limitations. The model assumes homogeneous mixing and no cross-cluster transmission; real hospitals have heterogeneous contact networks. Only 6 clusters were simulated (3 per arm). Drug B is modeled as fully effective against resistant strains; in practice, broad-spectrum agents may retain only partial activity, which would reduce the DE and attenuate the competitive exclusion tradeoff. The model omits de novo resistance emergence, patient heterogeneity, polymicrobial infections, and prior antibiotic exposure. Treatment allocation was deterministic; clinician behavior would be heterogeneous in reality. Our intent is not to predict specific clinical outcomes but to demonstrate, as a proof of principle, that the 2SR framework can detect and quantify spillover—arising from interference between patients’ treatment assignments—that conventional individually randomized designs cannot identify. Additionally, in our simulation, the infection status, strain type, treatment assignment, and transmission events of every patient are known at every time step—an omniscience that is impossible in practice. In a real trial, untreated infected patients may go unidentified, total deaths are difficult to attribute to specific strain exposures, and the encounters between susceptible and resistant infections that drive competitive exclusion are unobservable. This simplification serves the proof-of-principle aim, but substantial methodological work remains for both the design and analysis of a real 2SR trial and for the emulation of target trials with observational data. Many variations of the agent-based model could be developed to explore alternative scenarios (e.g., partial Drug B resistance, multiple strains, heterogeneous contact patterns), but the core contribution is methodological: establishing that the 2SR design is informative for antimicrobial evaluation.

This work establishes, as a proof of principle, that the 2SR design paired with ABM emulation can quantify both direct and indirect effects of antimicrobial strategies in the presence of transmission dynamics and resistance. A practical next step would be to implement a 2SR trial in a hospital system to determine whether population-level prescribing changes achieve the intended goal of reducing resistance without shifting disease burden to other patients.

## Data Availability

All data produced in the present study and the code for the simulation are available upon reasonable request to the authors

## Acknowledgments

The authors acknowledge support from the VK Fund for the CCDD.

---

**eFigure 1.**
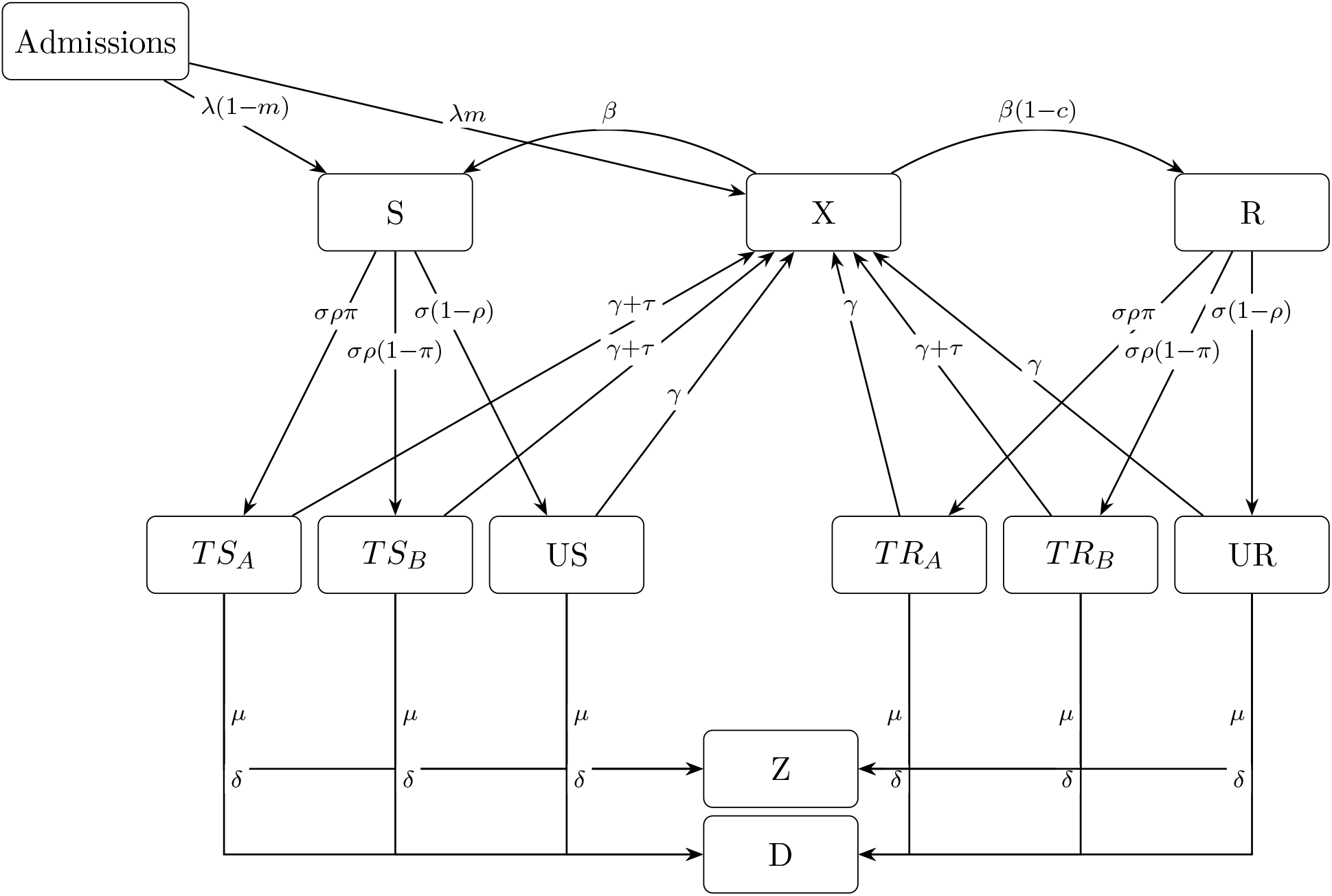
State-Transition Diagram. State-transition diagram for the hospital transmission ABM. Agents transition between uninfected (*X*), colonized (*S, R*), symptomatic/treated (*TS*_*A*_, *TS*_*B*_, *TR*_*A*_, *TR*_*B*_), symptomatic/untreated (*US, UR*), discharged (*Z*), and deceased (*D*) states. Drug A covers only susceptible strains; Drug B covers both.

**eFigure 2.**
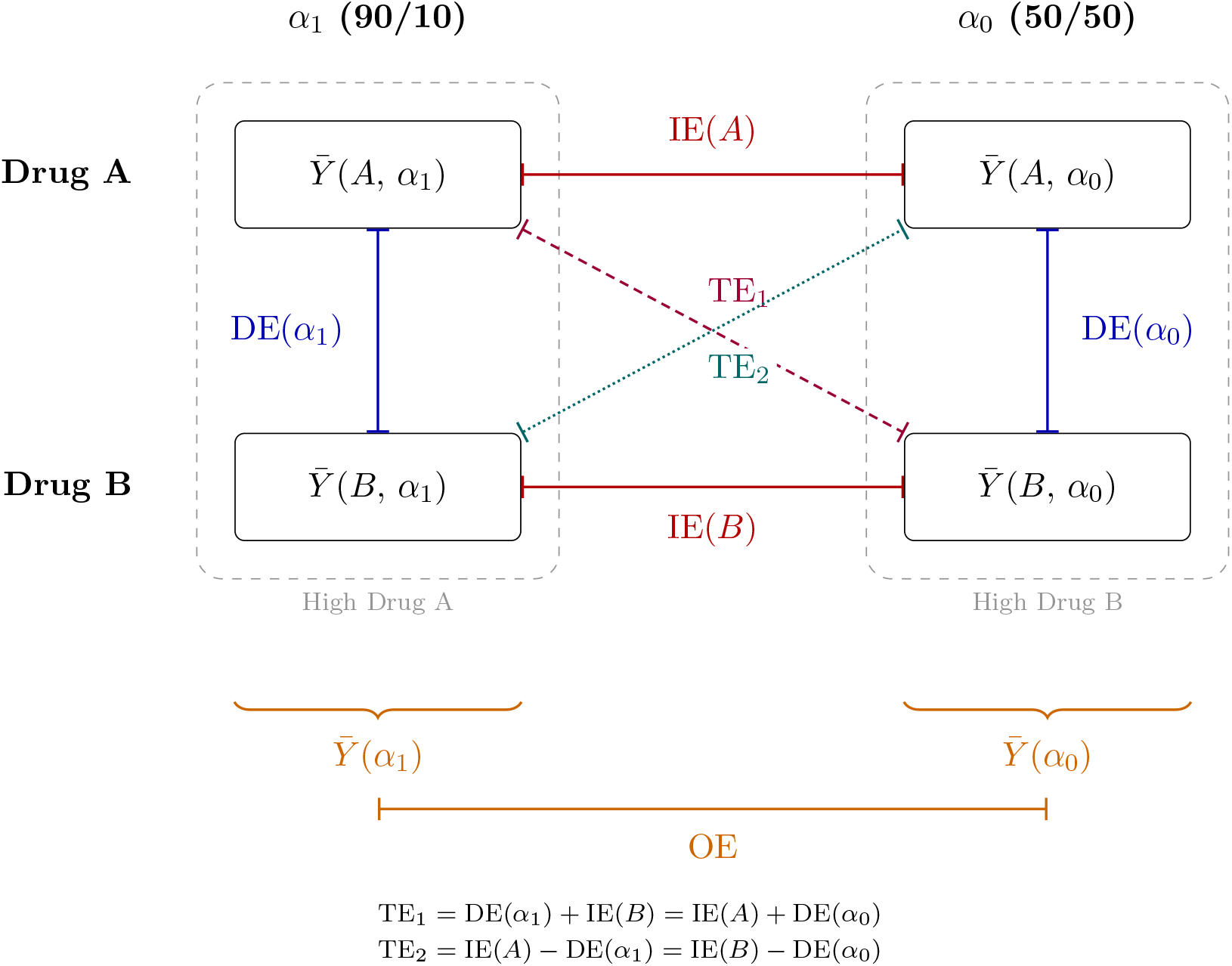
Causal Effect Decomposition. Causal effect decomposition under the two-stage randomization design, adapted from Hudgens and Halloran. ^2^ Each node represents a population-average potential outcome 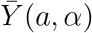 under treatment *a* ∈ {*A, B*} and allocation strategy *α*. Lines indicate contrasts (not causal pathways). Blue: direct effects (DE), comparing Drug A vs. Drug B within a strategy. Red: indirect effects (IE), comparing strategies for the same drug. Purple dashed: total effect 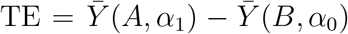. Teal dotted: second total effect 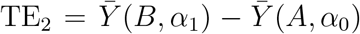, which arises because both treatments are active (in vaccine trials with a placebo arm, only one diagonal is natural). Orange: overall effect (OE), comparing marginal outcomes across strategies. The dashed ovals group the two strategy environments.

**eTable 1.**
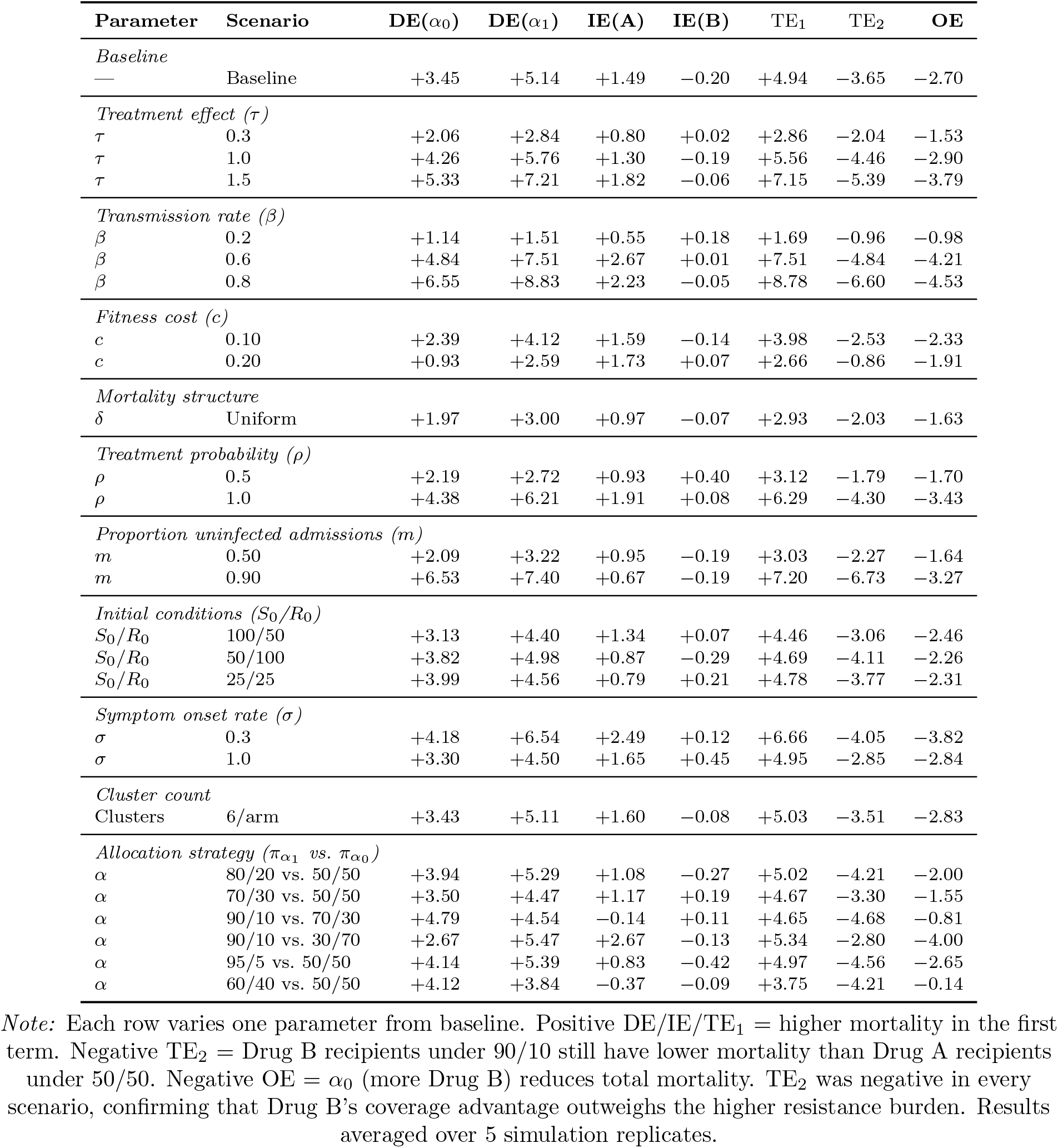
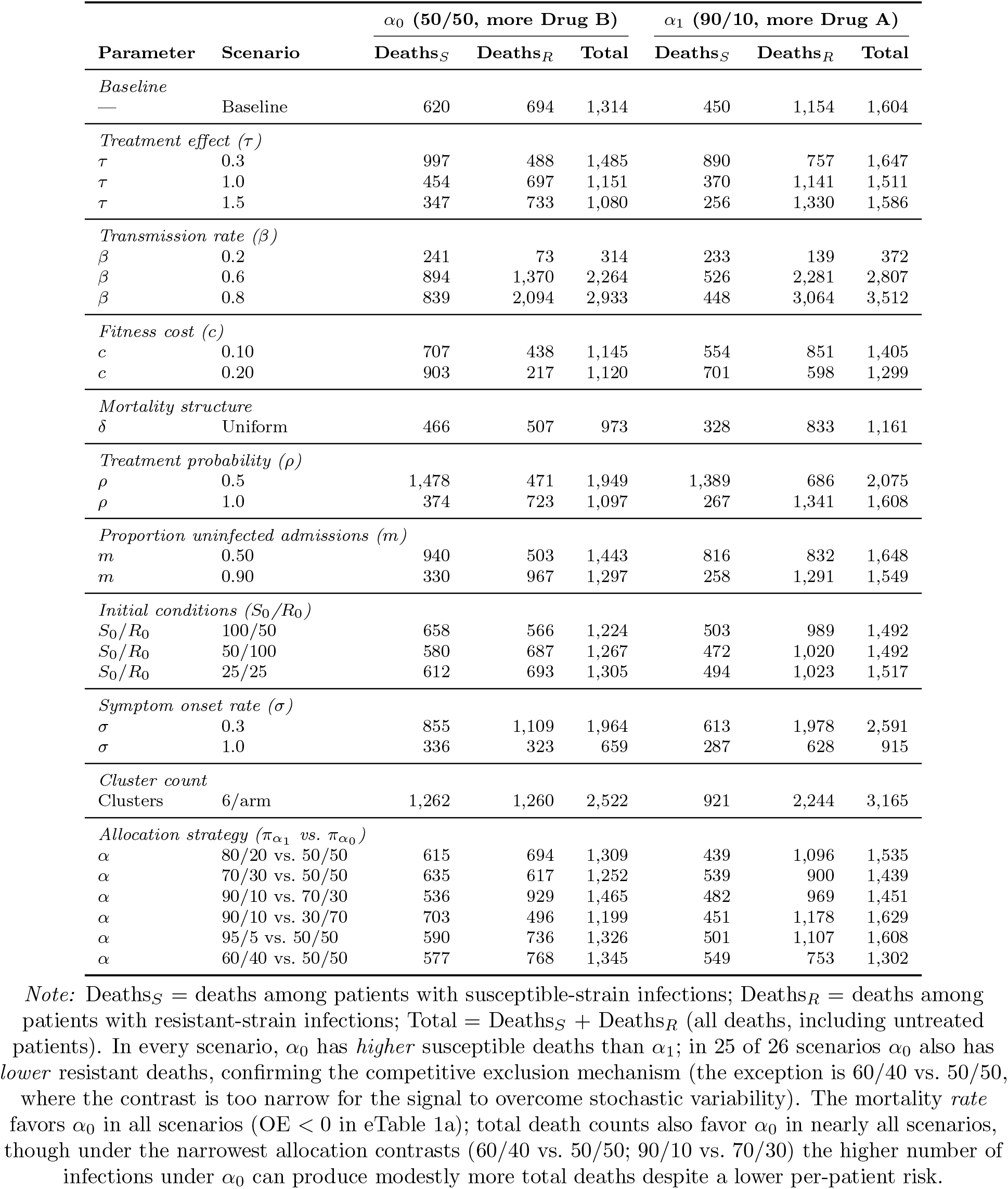
Comprehensive Sensitivity Analysis. **eTable 1a**. Causal effect estimates (percentage points) across sensitivity scenarios. DE = direct effect; IE = indirect effect; TE_1_, TE_2_ = total effects; OE = overall effect. *α*_0_ = low-Drug-A strategy (50/50); *α*_1_ = high-Drug-A strategy (90/10). Baseline: *τ* = 0.7, *β* = 0.4, *c* = 0.01, *σ* = 0.5, differential mortality, *ρ* = 0.9, *m* = 0.75, *S*_0_ = *R*_0_ = 50, 3 clusters/arm. **eTable 1b**. Deaths by strain type and allocation strategy, demonstrating the competitive exclusion tradeoff. Under *α*_0_ (50/50, more Drug B), resistant-strain deaths decrease but susceptible-strain deaths *increase* relative to *α*_1_ (90/10, more Drug A). This paradoxical redistribution occurs because Drug B suppresses resistant strains, freeing ecological space for susceptible-strain transmission. The mortality rate (OE) favors *α*_0_ in every scenario.

**eTable 2.**
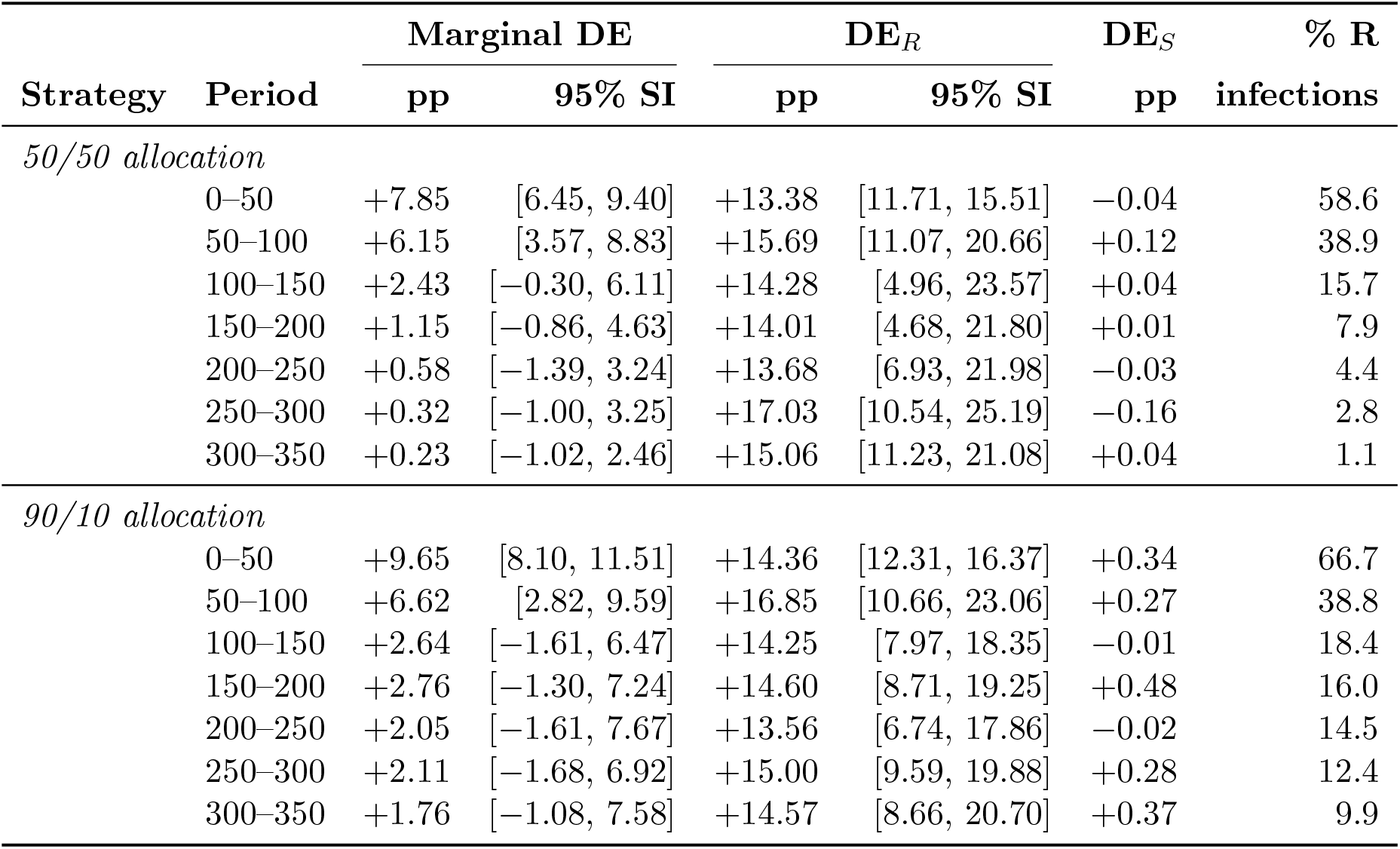
Time-Varying Direct Effects. Period-specific direct effects (DE) by allocation strategy, computed in 50-timestep windows. The marginal DE declines over time as the proportion of resistant infections decreases. Stratum-specific effects remain stable: DE_*S*_ ≈ 0 and DE_*R*_ ≈ 14 pp where estimable. DE_*R*_ is not reported (—) when fewer than 5 replicates have ≥ 30 resistant infections per drug arm. Values are means across 100 simulation replicates; 95% SI = 95% simulation interval.

